## Supplementary materials for "Relative income and its relationship with mental health in UK employees: a conceptual and methodological review"

S1. PRISMA Checklist

| **Section and Topic** | **Item #** | **Checklist item** | **Location where item is reported** |
| --- | --- | --- | --- |
| **TITLE** | | |  |
| Title | 1 | Identify the report as a systematic review. | Page 1 |
| **ABSTRACT** | | |  |
| Abstract | 2 | See the PRISMA 2020 for Abstracts checklist. | Page 2-3 |
| **INTRODUCTION** | | |  |
| Rationale | 3 | Describe the rationale for the review in the context of existing knowledge. | Pages 4-5 |
| Objectives | 4 | Provide an explicit statement of the objective(s) or question(s) the review addresses. | Page 5 |
| **METHODS** | | |  |
| Eligibility criteria | 5 | Specify the inclusion and exclusion criteria for the review and how studies were grouped for the syntheses. | Page 7 |
| Information sources | 6 | Specify all databases, registers, websites, organisations, reference lists and other sources searched or consulted to identify studies. Specify the date when each source was last searched or consulted. | Page 6 |
| Search strategy | 7 | Present the full search strategies for all databases, registers and websites, including any filters and limits used. | Supp 1 |
| Selection process | 8 | Specify the methods used to decide whether a study met the inclusion criteria of the review, including how many reviewers screened each record and each report retrieved, whether they worked independently, and if applicable, details of automation tools used in the process. | Pages 7-8 |
| Data collection process | 9 | Specify the methods used to collect data from reports, including how many reviewers collected data from each report, whether they worked independently, any processes for obtaining or confirming data from study investigators, and if applicable, details of automation tools used in the process. | Pages 7-8 |
| Data items | 10a | List and define all outcomes for which data were sought. Specify whether all results that were compatible with each outcome domain in each study were sought (e.g. for all measures, time points, analyses), and if not, the methods used to decide which results to collect. | Pages 7-8 |
|  | 10b | List and define all other variables for which data were sought (e.g. participant and intervention characteristics, funding sources). Describe any assumptions made about any missing or unclear information. | Page 8 |
| Study risk of bias assessment | 11 | Specify the methods used to assess risk of bias in the included studies, including details of the tool(s) used, how many reviewers assessed each study and whether they worked independently, and if applicable, details of automation tools used in the process. | Page 9 |
| Effect measures | 12 | Specify for each outcome the effect measure(s) (e.g. risk ratio, mean difference) used in the synthesis or presentation of results. | Page 9 |
| Synthesis methods | 13a | Describe the processes used to decide which studies were eligible for each synthesis (e.g. tabulating the study intervention characteristics and comparing against the planned groups for each synthesis (item #5)). | Pages 9-10 |
|  | 13b | Describe any methods required to prepare the data for presentation or synthesis, such as handling of missing summary statistics, or data conversions. | Pages 9-10 |
|  | 13c | Describe any methods used to tabulate or visually display results of individual studies and syntheses. | N/A |
|  | 13d | Describe any methods used to synthesize results and provide a rationale for the choice(s). If meta-analysis was performed, describe the model(s), method(s) to identify the presence and extent of statistical heterogeneity, and software package(s) used. | N/A |
|  | 13e | Describe any methods used to explore possible causes of heterogeneity among study results (e.g. subgroup analysis, meta-regression). | N/A |
|  | 13f | Describe any sensitivity analyses conducted to assess robustness of the synthesized results. | N/A |
| Reporting bias assessment | 14 | Describe any methods used to assess risk of bias due to missing results in a synthesis (arising from reporting biases). | Page 10 |
| Certainty assessment | 15 | Describe any methods used to assess certainty (or confidence) in the body of evidence for an outcome. | N/A |
| **RESULTS** | | |  |
| Study selection | 16a | Describe the results of the search and selection process, from the number of records identified in the search to the number of studies included in the review, ideally using a flow diagram. | Page 11 & Fig 1 |
|  | 16b | Cite studies that might appear to meet the inclusion criteria, but which were excluded, and explain why they were excluded. | Page 20 |
| Study characteristics | 17 | Cite each included study and present its characteristics. | Page 11, Tables 1 & 2 |
| Risk of bias in studies | 18 | Present assessments of risk of bias for each included study. | Page 11,  Supp 4 |
| Results of individual studies | 19 | For all outcomes, present, for each study: (a) summary statistics for each group (where appropriate) and (b) an effect estimate and its precision (e.g. confidence/credible interval), ideally using structured tables or plots. | Table 1 & Supp 2 |
| Results of syntheses | 20a | For each synthesis, briefly summarise the characteristics and risk of bias among contributing studies. | Page 11 |
|  | 20b | Present results of all statistical syntheses conducted. If meta-analysis was done, present for each the summary estimate and its precision (e.g. confidence/credible interval) and measures of statistical heterogeneity. If comparing groups, describe the direction of the effect. | N/A |
|  | 20c | Present results of all investigations of possible causes of heterogeneity among study results. | N/A |
|  | 20d | Present results of all sensitivity analyses conducted to assess the robustness of the synthesized results. | N/A |
| Reporting biases | 21 | Present assessments of risk of bias due to missing results (arising from reporting biases) for each synthesis assessed. | Page 11 |
| Certainty of evidence | 22 | Present assessments of certainty (or confidence) in the body of evidence for each outcome assessed. | N/A |
| **DISCUSSION** | | |  |
| Discussion | 23a | Provide a general interpretation of the results in the context of other evidence. | Page 17-20 |
|  | 23b | Discuss any limitations of the evidence included in the review. | Page 20 |
|  | 23c | Discuss any limitations of the review processes used. | Page 20-21 |
|  | 23d | Discuss implications of the results for practice, policy, and future research. | Pages 21-22 |
| **OTHER INFORMATION** | | |  |
| Registration and protocol | 24a | Provide registration information for the review, including register name and registration number, or state that the review was not registered. | Page 6 |
|  | 24b | Indicate where the review protocol can be accessed, or state that a protocol was not prepared. | Page 6 |
|  | 24c | Describe and explain any amendments to information provided at registration or in the protocol. | Page 20 |
| Support | 25 | Describe sources of financial or non-financial support for the review, and the role of the funders or sponsors in the review. | Page 31 |
| Competing interests | 26 | Declare any competing interests of review authors. | Page 31 |
| Availability of data, code and other materials | 27 | Report which of the following are publicly available and where they can be found: template data collection forms; data extracted from included studies; data used for all analyses; analytic code; any other materials used in the review. | Available from corresponding author on request |

*From:*  Page MJ, McKenzie JE, Bossuyt PM, Boutron I, Hoffmann TC, Mulrow CD, et al. The PRISMA 2020 statement: an updated guideline for reporting systematic reviews. BMJ 2021;372:n71. doi: 10.1136/bmj.n71

For more information, visit: <http://www.prisma-statement.org/>

S2. Search strategy

The following electronic databases will be searched with MeSH and free-text terms adjusted appropriately for each database: PubMed (including MEDLINE and PubMed Central), PsycINFO, Scopus, Web of Science, Global Health, JSTOR, Business Source Complete (EBSCO), ScienceDirect and Emerald.

The reference lists of all relevant papers will be hand-searched to identify additional studies. GoogleScholar will be used to search for recent citations of included studies since they were published.

In addition, a search of grey literature will be conducted via consultation with experts, google search, databases such as OpenGrey and search of key organisation websites.

### General search strategy

The following search strategy will be used based on these terms:

| **Key word** | **Terms** |
| --- | --- |
| Relative Income | **“Relative Income”** |
| Mental Health | **“Mental Health”**  **OR**  **“Mental health problem”**  **OR**  **“Mental health condition”**  **OR**  **“Mental health disorder”**  **OR**  **“Mental ill-health”**  **OR**  **“Mental illness”**  **OR**  **“Mental disorder”**  **OR**  **“Mental ill health”**  **OR**  **Distress**  **OR**  **Stress**  **OR**  **Wellbeing**  **OR**  **Well-being**  **OR**  **Happiness**  **OR**  **Satisfaction**  **OR**  **“quality of life”**  **OR**  **Health (title, ab)** |
| Alcohol Misuse | **Alcohol**  **OR**  **"alcohol misuse"**  **OR**  **"alcohol dependence*"**  **OR**  **"alcohol use disorder"**  **OR**  **AUD**  **OR**  **alcoholism**  **OR**  **alcoholic**  **OR**  **drink*** |
| Working Population | **Worker***  **OR**  **Staff**  **OR**  **Workforce**  **OR**  **“working population”**  **OR**  **“work force”**  **OR**  **Personnel**  **OR**  **“Labo$r force”**  **OR**  **Employee***  **OR**  **(“working age” AND adults)** |

### Specific database search strategy

| **Database** | **Terms used** | **Filters Used** |
| --- | --- | --- |
| Medline (Ovid) | All above terms for mental health, alcohol AND “relative income”  Working population terms not used in search but will be used as exclusion criteria | English Language |
| JSTOR | ("mental health" OR "mental illness" OR happiness OR satisfaction OR wellbeing OR alcohol OR "alcohol misuse" OR "alcohol dependency") AND "relative income” AND  (Worker* OR Staff OR Workforce OR “working population”) | English Language |
| PubMed | All above terms for mental health, alcohol AND “relative income”  Working population terms not used in search but will be used as exclusion criteria | English Language |
| PsychINFO (Ovid) | All above terms for mental health, alcohol AND “relative income”  Working population terms not used in search but will be used as exclusion criteria | English Language |
| Global Health (Ovid) | All above terms for mental health, alcohol AND “relative income”  Working population terms not used in search but will be used as exclusion criteria | English Language |
| Science Direct | ("mental health" OR "mental illness" OR happiness OR satisfaction OR wellbeing OR alcohol OR "alcohol misuse" OR "alcohol dependency") AND "relative income"  Working population terms not used in search but will be used as exclusion criteria | English Language  Type: Review Articles, Research Articles and Book Sections |
| Web of Science | All above terms for mental health, alcohol AND “relative income”  Working population terms not used in search but will be used as exclusion criteria | English Language |
| Scopus | All above terms for mental health, alcohol AND “relative income”  Working population terms not used in search but will be used as exclusion criteria | English Language |
| Emerald | All above terms for mental health, alcohol AND “relative income”  EXCEPT “health”  Working population terms not used in search but will be used as exclusion criteria | English Language |
| Business Source Complete (EBSCO) | All above terms for mental health, alcohol AND “relative income”  Working population terms not used in search but will be used as exclusion criteria | English Language |

S4. List of included papers

1. Becchetti, L., Colcerasa, F., & Pisani, F. (2022). When income differences hurt or excite: The nonlinear effect of regional inequality on subjective wellbeing [Article]. *Review of Income & Wealth*, 1. <https://doi.org/10.1111/roiw.12608>
2. Becchetti, L., Corrado, L., & Rossetti, F. (2011). The Heterogeneous Effects of Income Changes on Happiness [Article]. *Social indicators research*, *104*(3), 387-406. <https://doi.org/10.1007/s11205-010-9750-0>
3. Blanchflower, D. G. & oswald, A. J. (2004). Well-being over time in Britain and the USA. *Journal of Public Economics,* 88**,** 1359.
4. Brown, S., Gray, D., & Roberts, J. (2015). The relative income hypothesis: A comparison of methods [Article]. *Economics Letters*, *130*, 47-50. <https://doi.org/10.1016/j.econlet.2015.02.031>
5. Davidson, R., Kitzinger, J., & Hunt, K. (2006). The wealthy get healthy, the poor get poorly? Lay perceptions of health inequalities. *Social Science & Medicine*, *62*(9), 2171-2182. [https://doi.org/https://doi.org/10.1016/j.socscimed.2005.10.010](https://doi.org/https:/doi.org/10.1016/j.socscimed.2005.10.010)
6. FitzRoy, F. R., & Nolan, M. A. (2022). Income Status and Life Satisfaction [Article]. *Journal of happiness studies*, *23*(1), 233-256. <https://doi.org/10.1007/s10902-021-00397-y>
7. FitzRoy, F. R., Nolan, M. A., Steinhardt, M. F., & Ulph, D. (2014). Testing the tunnel effect: comparison, age and happiness in UK and German panels [Article]. *IZA Journal of European Labor Studies*, *3*(1), Article 24. <https://doi.org/10.1186/2193-9012-3-24>
8. Francis-Devine, B. (2022). *Poverty in the UK: statistics* (7096). <https://researchbriefings.files.parliament.uk/documents/SN07096/SN07096.pdf>
9. Fumagalli, E., & Fumagalli, L. (2022). Subjective well-being and the gender composition of the reference group: Evidence from a survey experiment. *Journal of Economic Behavior & Organization*, *194*, 196-219. [https://doi.org/https://doi.org/10.1016/j.jebo.2021.12.016](https://doi.org/https:/doi.org/10.1016/j.jebo.2021.12.016)
10. Lorgelly, P. K., & Lindley, J. (2008). What is the relationship between income inequality and health? Evidence from the BHPS. *Health Econ*, *17*(2), 249-265. <https://doi.org/10.1002/hec.1254>
11. Parker, L., Watson, D., & Webb, R. (2011). Family fortunes: Gender-based differences in the impact of employment and home characteristics on satisfaction levels. *The Journal of Socio-Economics*, *40*(3), 259-264. [https://doi.org/https://doi.org/10.1016/j.socec.2011.01.009](https://doi.org/https:/doi.org/10.1016/j.socec.2011.01.009)
12. Theodossiou, I., & Zangelidis, A. (2009). The social gradient in health: The effect of absolute income and subjective social status assessment on the individual's health in Europe. *Economics & Human Biology*, *7*(2), 229-237. [https://doi.org/https://doi.org/10.1016/j.ehb.2009.05.001](https://doi.org/https:/doi.org/10.1016/j.ehb.2009.05.001)
13. Yu, H. (2019). The Impact of Self‐Perceived Relative Income on Life Satisfaction: Evidence from British Panel Data [Article]. *SOUTHERN ECONOMIC JOURNAL*, *86*(2), 726-745. <https://doi.org/10.1002/soej.12349>
